# Effects of mindfulness meditation and Acceptance and Commitment Therapy in patients with obstructive sleep apnea with residual excessive sleepiness: A randomized controlled pilot study

**DOI:** 10.1101/2022.12.14.22283432

**Authors:** Max Hellrigel-Holderbaum, Nina Romanczuk-Seiferth, Martin Glos, Ingo Fietze

**Affiliations:** Interdisciplinary Center of Sleep Medicine, Charité – Universitätsmedizin Berlin, Berlin, Germany; Berlin School of Mind and Brain, Humboldt-Universität zu Berlin, Berlin, Germany; Department of Psychiatry and Psychotherapy, Charité – Universitätsmedizin Berlin, Berlin, Germany; The Fourth People’s hospital of Guangyuan, Guangyuan City, China

## Abstract

**Objective:** Assessing the effects of Mindfulness-Based Stress Reduction (MBSR) and Acceptance and Commitment Therapy (ACT) on symptoms of OSA, especially on the main outcome, excessive daytime sleepiness (EDS).

**Methods:** Parallel randomized controlled trial. 16 OSA patients with residual EDS (rEDS) were randomized and assigned to the two programs via a central computer system using REDCap. Participants completed a standardized 8-week MBSR program or a time-matched program on Acceptance and Commitment Therapy (ACT). Both programs were conducted online. Participants answered questionnaires online at baseline (pre), post-intervention (post), three months after the intervention (follow-up) and were blinded to whether their group was the treatment or control group but not to group allocation. Three participants dropped out early. Most analyses are based on the remaining 13 patients.

**Results:** There was a significant difference between the MBSR (n=7) and ACT group (n=6) in changes of EDS between pre and post (p=.043) and a significant reduction of EDS for patients in the ACT group at post (p=.034). This EDS reduction averaging 2.17 points on the Epworth Sleepiness Scale reached the prespecified bar for clinical significance of two points on that scale. Insomnia symptoms reduced significantly following ACT (p=.017). In MBSR, both participants and the MBSR-trainer judged movement-based exercises to be most efficacious.

**Conclusion:** ACT shows potential as adjunctive therapy for OSA with rEDS, although further studies are needed. It seems promising to develop therapeutic approaches for OSA with rEDS using ACT, especially if they are tailored to patients’ needs.

**Trial registration:** drks.de; Identifier: DRKS00026812

## 1. Introduction

Obstructive sleep apnea (OSA) is a highly prevalent sleep disorder that affects approximately 25% of adults in the US and similar amounts in other high-income countries, although estimates vary depending on its definition (Gottlieb & Punjabi, 2020; Benjafield et al., 2019). Excessive daytime sleepiness (EDS) is one of the most important clinical symptoms of OSA and is often treated effectively with positive airway pressure (PAP) therapy (Fietze et al., 2021; Gottlieb & Punjabi, 2020). Without treatment, OSA is associated especially with cardiovascular comorbidities as well as cognitive and executive problems (Fietze et al., 2021; Lévy et al., 2015). Patients further suffer from a reduced quality of life (Jehan et al., 2017). Despite PAP therapy, at least 10% of patients still suffer from residual EDS (rEDS) (Fietze et al., 2021; Gasa et al., 2013; Javaheri & Javaheri, 2020; Launois et al., 2013; Pépin et al., 2009). In the largest study until now, using data from 1047 patients, a total prevalence of 13% of rEDS in OSA patients was estimated (Gasa et al., 2013). There are other treatment options for patients with OSA besides the gold standard of PAP therapy, including most importantly pharmacotherapy in the case of rEDS after primary therapy (Fietze et al., 2021). Pharmacotherapeutic approaches, i.e. especially Solriamfetol in the US and Europe, reduce daytime sleepiness but often come with side effects such as headache, nausea, decreased appetite, and anxiety (Schweitzer et a., 2019). At the same time, behavioral therapies remain largely unexplored, resulting in a lack of behavioral therapies established in practice for sleep disorders in general, including OSA (Pigeon et al., 2007). Cognitive Behavioral Therapy for Insomnia (CBT-I) is an encouraging exception. CBT-I showed a similar short-term and better long-term efficacy than established pharmacotherapy, and has subsequently become the recommended first-line treatment for primary insomnia (OKAJIMA et al., 2011; Riemann & Perlis, 2009; Trauer et al., 2015; van Straten et al., 2018).

Mindfulness is a non-judgmental form of attention of the present moment, often trained through meditation (Kabat-Zinn, 2015; Shapiro et al., 2006). Research on mindfulness meditation (MM) has grown substantially in the last 15 years (Creswell, 2017; Goleman & Davidson, 2018). Mindfulness-Based Stress Reduction (MBSR) is the most thoroughly studied program for training MM. Note that while MM is often seen as a mere relaxation exercise, traditionally, MM is rather described as avoiding strong agitation *and* drowsiness (Britton et al., 2014). Studies on MM and MBSR found various improvements in both clinical populations and the general population. Some of the most notable improvements include chronic pain, depressive symptoms, depression relapse, anxiety symptoms, addictive disorders, and quality of life (Creswell, 2017; Vibe et al., 2017). Studies in sleep medicine found MM to improve sleep quality (Black et al., 2015; Garland et al., 2014; Rash et al., 2019; Rusch et al., 2019) and sleep disturbance (Huberty et al., 2021; Kanen et al., 2015; Nakamura et al., 2013). While these results show the efficacy of MM, its *comparative effectiveness* to established treatments, especially regarding anxiety, quality of life, sleep quality and sleep disturbance is much less clear and often shows mixed results or insufficient evidence (Goyal et al., 2014; Vibe et al., 2017; Black et al., 2015; Rash et al., 2019; Rusch et al., 2019; Huberty et al., 2021; Kanen et al., 2015). Studies with stronger designs are needed to judge the comparative effectiveness of MM and MBSR (van Dam et al., 2018).

Earlier phases in training MM tend to be associated with improvements in orienting and executive attention while later phases tend to be associated with alerting attention (Chiesa et al., 2011; Tang et al., 2015). While more research is needed to consolidate these findings, they have not yet been studied in patients with EDS or specifically OSA patients with rEDS. MM and MBSR were shown to reduce fatigue symptoms in clinical groups like breast cancer patients and the general population (Xie et al., 2020; Zeidan et al., 2010; Zhang et al., 2019). Importantly, the symptoms of fatigue and EDS are similar, and in clinical practice often difficult to differentiate. In a recent study using a mobile app to deliver MM to patients with sleep disturbance, both daytime sleepiness and fatigue were reduced (Huberty et al., 2021). Notably though, the program was substantially less intense than the standard MBSR program in requiring only 10 mins/ day of meditation and the reported effect was rather small (less than one point difference on the ESS) and thus not clinically significant. Thus, the effects of an intensive course on daytime sleepiness remain open.

Acceptance- and Commitment Therapy (ACT) is a behavioral therapy of the third generation, i.e., it leans on principles of behavioral therapy, including cognitive behavioral therapy, but has developed these further (Hayes, 2004). ACT focuses on the development of psychological flexibility which involves defusion and acceptance of thoughts, a contextual sense of self, and commitment to personal values in everyday behavior (Hayes et al., 2006). Its effectiveness in treating depression, anxiety disorders, substance use disorders, and chronic pain has been shown in several meta-analyses (Gloster et al., 2020). In sleep medicine, it has recently been shown to reduce symptoms of insomnia, thereby showing potential for an adjunctive behavioral therapy besides CBT-I (Paulos-Guarnieri et al., 2022; Salari et al., 2020).

In this study, we wanted to investigate whether some of the findings on MM would translate to patients with OSA and rEDS. We aimed to investigate the effects of both MBSR and ACT on core variables of interest, including especially daytime sleepiness, quality of life, sleep quality and sleep disturbance in these patients. We also aimed to test the online format and different components of both interventions. As our primary hypothesis, we expected MBSR to reduce EDS for patients with OSA and rEDS. We expected this effect to be clinically significant, that is on average exceed 2 points on the ESS in the MBSR group after the program. This expectation was primarily based on the aforementioned effects of MM and MBSR on fatigue (Zeidan et al., 2010; Zhang et al., 2019). Further, the effects of MM on alerting attention (Chiesa et al., 2011; Tang et al., 2015), sleep quality (Rusch et al., 2019), and the negative correlation between mindfulness and daytime sleepiness (Howell et al., 2010) supported this hypothesis. Second, we expected MBSR to reduce EDS for OSA patients with rEDS beyond the active control group engaging in ACT group sessions. This hypothesis drew equally from previous results while we were not aware of similar findings for ACT. We further expected MBSR to improve sleep quality and reduce sleep disturbance for our patients beyond the active control group. Finally, we hypothesized MBSR to raise the participants’ HRQoL beyond the (specific) active control.

## 2. Methods

The study was conducted as an online study using REDCap (Research Electronic Data Capture) (Harris et al., 2009; Harris et al., 2019) tools hosted at Charité – Universitätsmedizin Berlin to gather data online via questionnaires. Video conferencing was used to conduct the group sessions. We ran the interventions from October 2021 to December 2021.

### 2.1 Participants

Of the 16 patients who met the inclusion and exclusion criteria and were enrolled in the study, 13 completed the study, including the subsequent and 3-month follow-up survey. The participants were aged between 28 and 69 years (M = 55.38; SD = 10.91). See table 1 for an overview of the participants’ baseline characteristics. Four of them were female, reflecting the higher prevalence of men in OSA patients. Patients on average used their PAP device for 4.3 hours. We did not exclude patients that were non-adherent to the PAP-therapy (PAP adherence defined as a usage of ≥ 4 hours per night for at least 70% of the nights) since we saw these patients as equally apt for behavioral therapy. Nevertheless, most patients that were enrolled were PAP adherent at baseline (n = 11 or 69%). There was no significant group difference in the number of PAP adherent subjects (χ2 = 2.45, p = .12) using ‘N-1’ chi-squared test as recommended by (Campbell, 2007). Neither was a difference between the groups at baseline significant at an alpha level of .05 using independent t-tests. Participants were recruited through personal communication and flyers in doctor’s consultation of the outpatient department of the Interdisciplinary Center of Sleep Medicine at Charité-Universitätsmedizin Berlin. Further, potential participants were contacted via self-help groups focusing on OSA, including both local ones throughout Germany and online groups in social networks. To be included, only potential participants with a diagnosis of obstructive sleep apnea, an ESS score of above 10, and an age between 18 and 75 years were considered. Further, they needed an ongoing PAP therapy that started at least a month ago, or a previous attempt at a PAP therapy without starting another therapy of sleep apnea during the study. Moreover, only potential participants without an extensive previous experience with MM including MBSR, formal exercises for stress management, or ACT were considered. Patients with acute major depressive disorder or suicidality, acute or chronic conditions requiring ongoing treatment were excluded. Drug or alcohol dependency and insufficient knowledge of the German language were also used as exclusion criteria. Participants did not receive monetary reimbursement for their participation. The study was approved by the local ethics committee of the Charité-Universitätsmedizin Berlin (EA1/139/21) and conducted in accordance with the declaration of Helsinki. All participants provided informed written consent.

**Table 1:** Demographics and Clinical Characteristics of participants before intervention Baseline characteristics of the two groups and overall characteristics expressed as means ± standard deviation. Abbreviations: ESS, Epworth Sleepiness Scale; MBSR, mindfulness-based stress reduction; ACT, acceptance and commitment therapy; SD, standard deviation

|  | MBSR, intervention<br>(n=8) | ACT, control<br>(n=8) | Overall<br>(n=16) |
| --- | --- | --- | --- |
| Age (years) | 50.5 ± 12.8 | 60.3 ± 6.1 | 55.4 ± 10.9 |
| Sex (Number and Percentage of Females / Males) | 1 (12.5%) /<br>7 (87.5%) | 3 (37.5%) /<br>5 (62.5%) | 4 (25.0%) /<br>12 (75.0%) |
| Daytime sleepiness (ESS score) | 14.5 ± 3.8 | 14.1 ± 2.2 | 14.3 ± 3.0 |
| Usage of PAP device (hours per night) | 5.1 ± 2.2 | 3.4 ± 3.0 | 4.3 ± 2.7 |
| Usage of PAP device (days per week) | 5.9 ± 2.4 | 3.4 ± 3.0 | 4.6 ± 2.9 |
| BMI | 28.8 ± 5.8 | 26.4 ± 3.2 | 27.6 ± 4.7 |
| Weight (kg) | 88.6 ± 10.6 | 80.8 ± 13.7 | 84.7 ± 12.5 |
| Height (cm) | 176 ± 9.3 | 175 ± 7.9 | 175 ± 8.4 |
| Alcohol consumption (drinks per week) | 5.0 ± 5.1 | 1.4 ± 2.0 | 3.2 ± 4.2 |
| Interest in behavioral therapy for OSA symptoms (1-10) | 9.0 ± 1.2 | 8.0 ± 3.2 | 8.5 ± 2.4 |

### 2.2 Procedure

Potential participants were screened following the in- and exclusion criteria mentioned above and asked to fill out an ESS questionnaire that assessed whether they had an ESS score of above 10. They were subsequently informed about the detailed procedure and dates for participation in the study. Eligible participants were randomized to one of two groups. The first group (intervention group) engaged in a MBSR program as standardized by Kabat-Zinn and most recently Santorelli (Santorelli et al., 2017). This is the most studied program for mindfulness meditation which, besides classical sitting meditation, includes soft Yoga and stretching exercises, walking meditation, and body scans. These partially more physically active exercises of mindfulness were presumed to be especially appropriate for the participants with excessive daytime sleepiness, the idea being that soft body exercises would maintain basic alertness for training mindfulness and attention. A professional MBSR trainer with 10 years of experience in MBSR (teaching and practice) led the program.

The second group (active control group) completed a program focused on Acceptance and Commitment Therapy (ACT) theory and exercises. It aimed to develop psychological flexibility via several routes. These included perspective-taking, metaphors, and several experiential exercises, some of which focus on developing a less rigid and more contextual sense of self.

Another aim was to help participants align their everyday behavior with their values by clarifying and subsequently committing to them. A further aspect of this program could be described as stress management by changing the individual’s relationship to stressors after these have been recognized as such, for example via cognitive defusion. The program in part specifically addressed emotional difficulties associated with symptoms of sleep apnea. Finally, light stretching- and strengthening exercises comparable to the MBSR group were included. While ACT has been termed a mindfulness-related intervention (Creswell, 2017), we made sure to avoid exercises specifically designed for mindfulness as they are used in MM including MBSR. This program was led by a trained psychotherapist (N.R.-S.) with 10 years of experience with ACT (teaching and practice).

Both programs consisted of 8 online sessions with additional daily exercises of varying but similar lengths and a 6-hour retreat on a weekend towards the end of the program. Both programs were conducted by qualified and experienced professionals. After being randomized to one of the groups, participants completed a survey on demographics and clinical parameters of interest, elaborated below, with respective standardized questionnaires. Questionnaires were filled out before and after the completion of the programs as well as after a 3-months follow-up. An additional short questionnaire on participants’ expectations about their program was filled out after the second group session. Subsequently, the programs were conducted as described. In both programs, the teachers (independently) got sick for one week so the programs were extended for one week to make up for the missed session.

### 2.3 Materials

Before being enrolled, participants received a course plan with descriptions and timelines of the two programs, besides general information on the study. During the programs, participants received handouts to follow up on the group sessions that contained exercises and audio files with instructions for (daily) exercises.

#### Questionnaires

The Epworth sleepiness scale (ESS) (Johns, 1991) and Stanford Sleepiness Scale (SSS) (Hoddes et al., 1972) were used to measure daytime sleepiness. Variables concerning participants’ sleep, such as sleep quality, sleep latency, sleep efficiency, total sleep time and sleep disturbance, were assessed with the *Pittsburgh Sleep Quality Index* (PSQI) (Buysse et al., 1989). Health-related quality of life (HRQoL) concerning OSA symptoms, particularly EDS, was assessed with the *Functional Outcomes of Sleep Questionnaire* (FOSQ-10) (Chasens et al., 2009) and the general quality of life (QoL) was measured with the *Short Form 36* questionnaire (SF-36) (Ware & Sherbourne, 1992). To measure depressive symptoms, the *Patient Health questionnaire* (PHQ-9) (Kroenke et al., 2001) was used. The *Hospital Anxiety and Depression Scale* (HADS) (Zigmond & Snaith, 1983) was used to capture both depressive and anxious symptoms. Participants’ stress was measured with the *Perceived Stress Questionnaire* (PSQ) (Fliege et al., 2009). Insomnia symptoms were assessed with the *Insomnia Severity Index* (ISI) (Bastien, 2001). To control for the efficacy of the MBSR and ACT programs, participant’s mindfulness was assessed with the *Mindful Attention Awareness Scale* (MAAS) (K. W. Brown & Ryan, 2003) and psychological flexibility was measured with the *Acceptance and Action Questionnaire* (AAQ-II) (Bond et al., 2011). Finally, to gather quantitative data on participants’ perceived credibility and expectancy concerning their respective program, the *Credibility/ Expectancy Questionnaire* (CEQ) (Devilly & Borkovec, 2000) was used.

### 2.4 Design

The study was a parallel-group interventional pilot randomized controlled trial (trial registration: drks.de DRKS-ID: DRKS00026812). Participants’ assessments were conducted pre-intervention (i.e., at baseline), post-intervention (9 weeks), and follow-up (3 months).

The group engaging in ACT was chosen as an active control for EDS, other sleep-related variables, and HRQoL, thereby serving as an active control for all our hypotheses. We additionally looked at depressive and anxious symptoms, which are elevated in OSA. For these variables, MBSR was a specific active control condition of the ACT group since the effects of ACT are better established here. Thus, both programs served as controls for each other concerning different variables of interest. For the group comparisons in which the MBSR group served as the control condition, we did not set hypotheses since we did not expect to be able to find significant group differences given our sample size. The analyses on these variables are therefore exploratory. These crossed control conditions allowed us to explore the effects of both conditions on different variables with respectively strong control conditions.

We used ACT as a particularly strong active control condition for MBSR concerning EDS, HRQoL, and sleep variables due to the general need in the literature on MM/MBSR for these (Davidson & Kaszniak, 2015; Goyal et al., 2014; Kreplin et al., 2018) as well as for studies on MM and sleep quality in particular (Rash et al., 2019; Rusch et al., 2019).

We used block randomization with block sizes of 6, a 1:1 ratio, and a random assignment program using R version 4.0.5 and the blockrand package (Snow, 2020). The thereby created random allocation sequence was used in REDCap to allocate the participants to the groups.

The sample size was determined based on the recommendation of 20 participants for pilot studies with medium effect sizes (0.3 < Cohen’s d < 0.7) (Whitehead et al., 2016). The medium effect size was expected for the main outcome EDS based on a meta-analysis concerning the effects of MBSR on fatigue (Zhang et al., 2019). We failed to reach the goal of recruiting 20 participants before the beginning of the study and decided to run it with the recruited 16 participants as the teachers of the programs could not lead their courses later.

Partial blinding: Participants of the study were not blinded regarding which group (MBSR or ACT) they belong to. They were, however, blinded to which treatment group was the focus of the study and the different expected effects of each group as recommended by Davidson & Kaszniak (2015). To ensure this, we provided identical and general information to both groups regarding the potential positive effects (“improvement in symptoms of sleep apnea”). Partial blinding was employed to avoid different expectation effects as confounds of changes between the groups. PAP use, alcohol consumption and body weight were anticipated as extraneous variables and their potential influences tested below. Data were collected using automatically delivered online self-rating questionnaires. Groups were matched in the length of the intervention, amount of homework/practice, and the expertise of the instructors.

Regarding therapy setting, both programs were conducted online. We chose online delivery since several recent meta-analyses showed the efficacy of online delivery of ACT (M. Brown et al., 2016; Sommers-Spijkerman et al., 2021; Thompson et al., 2021) and a growing literature, including several meta-analyses, shows that online MM interventions show similar effects in improving common clinical outcomes of interest as in-person sessions (Sommers-Spijkerman et al., 2021; Spijkerman et al., 2016; Toivonen et al., 2017). Additionally, in other patient groups suffering from EDS, it was reported that patients preferred online instructions specifically because of their symptoms (Ong et al., 2020). Consequently, web-based programs for both MBSR and ACT were seen as an adequate method for instruction in our study while holding particular promise for scaling therapeutic approaches.

### 2.5 Statistical Analyses

All statistical analyses were performed in R version 4.0.5 (R Core Team, 2021). Descriptive statistics in Table 1 and Table 2 were obtained using the table1 package (Rich, 2021). To test potential differences at baseline between the two groups, ‘N-1’ chi-square tests, independent two-sample t-tests, and Welch two-sample t-tests were used. Paired samples t-tests were used to assess changes from baseline to post-treatment within groups. To compare between the groups, we used independent two-sample t-tests, comparing aggregate measures between groups. More specifically, we compared differences between the two time points between the groups (pre to post and pre to follow-up, and in case either of the former was significant, post to follow-up). This approach is one option for performing hypothesis tests in repeated measures designs with longitudinal data and works by condensing the data to one data point per subject, thus allowing for standard statistical hypothesis tests (Schober & Vetter, 2018).

**Table 2:** Characteristics of participants throughout the programs Characteristics of the two groups throughout the programs expressed as means ± standard deviation. Pre = before the intervention, Post = after the intervention, Follow-up = 3 months after Post. Abbreviations: MBSR, mindfulness-based stress reduction; ACT, acceptance and commitment therapy; SD, standard deviation

|  | Pre |  | Post |  | Follow-up |  |
| --- | --- | --- | --- | --- | --- | --- |
|  | MBSR, intervention (n=8) | ACT, control (n=8) | MBSR, intervention (n=7) | ACT, control (n=6) | MBSR, intervention (n=7) | ACT, control (n=6) |
| Daytime sleepiness (ESS score) | 14.5 ± 3.8 | 14.1 ± 2.2 | 16.0 ± 3.6 | 11.7 ± 1.6 | 14.4 ± 4.4 | 12.3 ± 2.9 |
| HRQoL (FOSQ score) | 13.6 ± 3.6 | 13.2 ± 3.4 | 13.2 ± 3.6 | 13.9 ± 2.4 | 13.2 ± 3.6 | 13.9 ± 2.4 |
| Sleep quality (PSQI score) | 8.6 ± 3.2 | 8.9 ± 3.5 | 7.7 ± 3.4 | 9.8 ± 2.6 | 8.1 ± 3.6 | 9.5 ± 3.0 |
| QoL (SF-36 score) | 55.1 ± 10.0 | 61.2 ± 13.3 | 60.0 ± 16.8 | 61.6 ± 15.3 | 56.2 ± 16.8 | 61.7 ± 11.2 |
| Usage of PAP device (hours per night) | 5.1 ± 2.2 | 3.4 ± 3.0 | 5.4 ± 2.5 | 3.3 ± 2.0 | 5.3 ± 2.4 | 3.8 ± 3.1 |
| Usage of PAP device (days per week) | 5.9 ± 2.4 | 3.4 ± 3.0 | 5.7 ± 2.6 | 4.0 ± 2.5 | 5.6 ± 2.5 | 3.3 ± 3.1 |
| Insomnia (ISI score) | 14.6 ± 4.4 | 15.9 ± 4.0 | 12.0 ± 5.9 | 15.3 ± 4.5 | 11.7 ± 7.6 | 15.5 ± 5.1 |
| Mindfulness (MAAS score) | 2.6 ± 1.0 | 3.8 ± 0.8 | 2.5 ± 0.6 | 3.7 ± 0.5 | 2.8 ± 0.8 | 3.6 ± 0.4 |
| Psychological flexibility (AAQ2 score) | 23.7 ± 7.6 | 21.4 ± 9.4 | 22.0 ± 7.9 | 18.5 ± 7.0 | 21.6 ± 10.3 | 17.7 ± 8.3 |
| Stress (PSQ score) | 60.2 ± 17.6 | 61.0 ± 16.6 | 51.7 ± 16.9 | 58.1 ± 21.3 | 52.9 ± 21.9 | 59.2 ± 20.7 |

In additional analyses, we used linear mixed-effects models (LMMs), which were fitted with full information maximum likelihood estimation and default convergence criteria. We added this more flexible design to our analyses primarily because of two important advantages. LMMs can use all available data for each subject while also handling missing data more appropriately (Brown, 2021; Gueorguieva & Krystal, 2004). Additional data points that were used in these models included both early data points from subjects that eventually dropped out as well as data (ESS scores) from screening. LMMs also allow for analyses with continuous dependent variables. For the implementation of LMMs, we used the lme4 package (Bates et al., 2015) and afex package (Singmann et al., 2021). In the LMMs, we found no obvious violations of homoscedasticity or normality using residual plots. When both models for model comparisons could be estimated, we reported model comparisons using likelihood-ratio tests and the summary of the mixed-model; otherwise, we only report the summary of the mixed-model.

## 3. Results

The 16 participants were randomized resulting in 8 participants in each of the two groups. Participants reported after the study to have taken part in on average 7.92 of the 9 sessions (SD = 1.04) while on average performing 48% of the daily exercises (SD = 21.45). These values were similar across groups and did not differ significantly from each other using independent t-tests. 3 participants in total dropped out of the study (i.e., they did not continue participation and did not complete post and follow-up surveys). All dropouts happened early (i.e., within the first 3 weeks), while there was no further dropout after that including the follow-up survey. 2 participants dropped out in the ACT group, 1 in the MBSR group. There was no significant difference in dropouts between the groups (χ2 = .38, df = 1, p = .5) using ‘N-1’ chi-squared test as recommended by (Campbell, 2007). There were also no differences in age (t = 1.05, df = 6.80, p = .33), PAP device usage in h per night (t = -1.31, df = 2.46, p = .30), PAP device usage in days per week (t = -1.52, df = 2.54, p = .24), sleepiness (t = -.41, df = 2.28, p = .72), HRQoL (t = .35, df = 1.19, p = .78), QoL (t = -.04, df = 1.17, p = .97), and sleep quality (t = -.61, df = 3.49, p = .58) when comparing participants who dropped out to those who remained in the study. Most analyses except the LMMs exclude the three dropouts.

### Daytime sleepiness (primary outcome)

Changes in daytime sleepiness from before to after the intervention as measured in ESS scores differed significantly between the MBSR group (*M* = .86, *SD* = 2.85) and ACT group (*M* = -2.17, *SD* = 1.83), *t*(10.3) = 2.30, *p* = 0.043 using a Welch two sample independent t-test (see Figure 1). The positive mean score in the MBSR group indicates a slight increase while the negative mean score of -2.17 in the ACT group indicates a reduction that surpassed our threshold for clinical significance of 2 points on the ESS from the preregistration. A large effect size was found with Cohen’s d = 1.24 though with a notably large 95% CI [0.01, 2,42]. Changes in daytime sleepiness however did not differ significantly from pre intervention to follow-up between the MBSR group (*M* = -.71, *SD* = 4.31) and ACT group (*M* = -1.50, *SD* = 3.02), *t*(10.6) = .38, *p* = .708 and from post intervention to follow-up between the MBSR group (*M* = 1.57, *SD* = 2.76) and ACT group (*M* = -0.67, *SD* = 3.93), *t*(8.8) = 1.17, *p* = .273. Follow-up analyses within the two groups using dependent paired samples t-tests confirmed a significant reduction in the ACT group from pre (*M* = 14.13, *SD* = 2.23) to post (*M* = 11.67, *SD* = 1.63), *t*(5) = 2.89, *p* = .034 while the daytime sleepiness did not change significantly in the MBSR group from before (*M* = 14.50, *SD* = 3.82) to after the intervention (*M* = 16.00, *SD* = 3.61), *t*(6) = -0.79, *p* = .457. Here, the effect size for the ACT group was similarly large with Cohen’s d_z_ = 1.18 and [0.08, 2.22] as 95% CI. A LMM in which we used additional data from the screening time point and data from dropouts, supported these findings. We used group, time, and the interaction between group and time as fixed effects and intercepts for subjects as random effects. While no main effect of time or group was significant, the interaction of (ACT) group and the post-treatment time point was marginally significant β = - 3.02, *SE* = 1.54, 95% CI [-5.90, -.16], *t*(34.2) = -1.96, *p* = .058.

**Figure 1.**
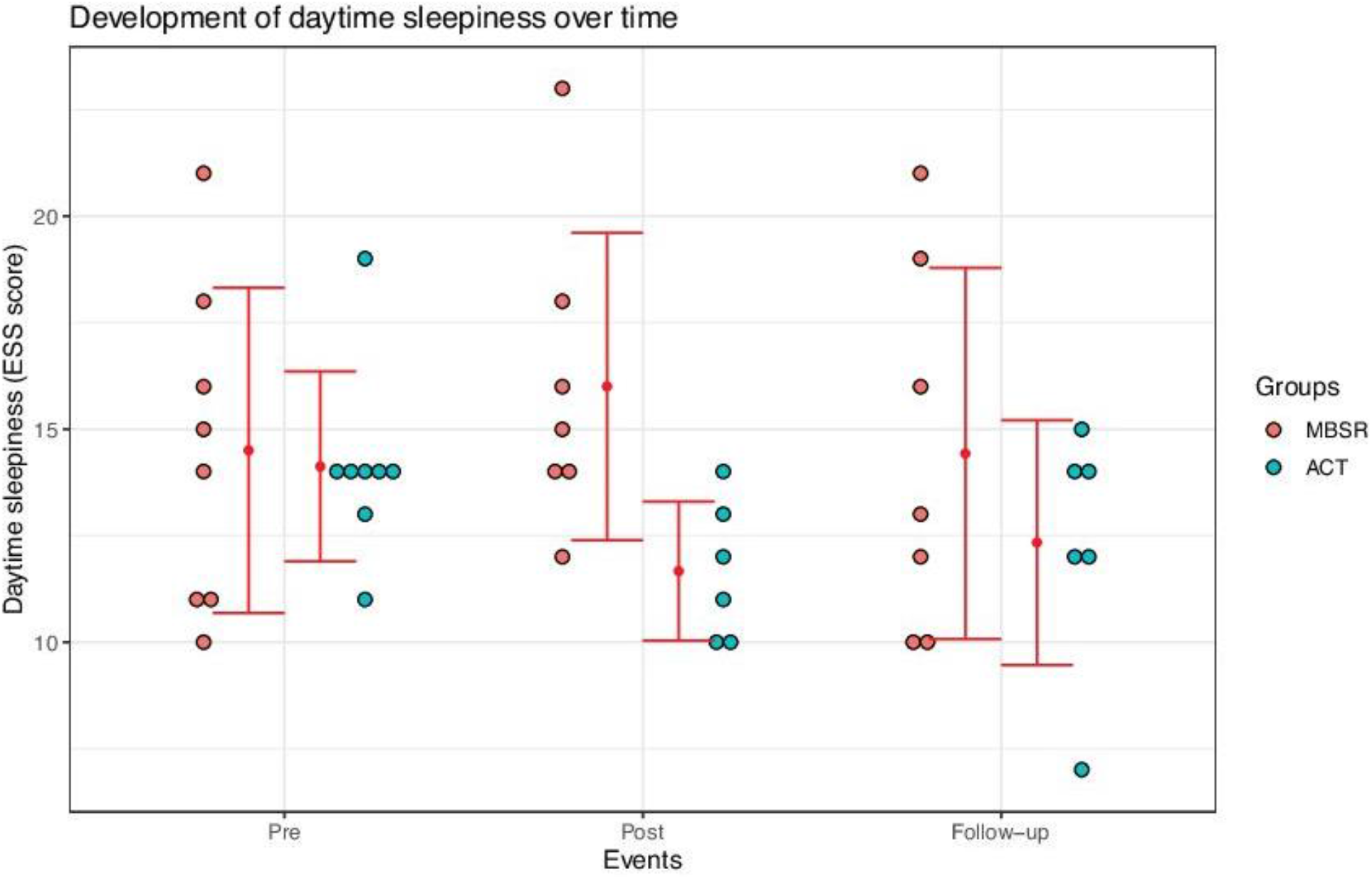
Development of daytime sleepiness throughout the study. ESS = Epworth Sleepiness Scale, bars represent ± one standard deviation, small red dots represent means.

### Secondary outcomes

Changes in HRQoL from pre to post differed marginally significantly between the MBSR group (*M* = -0.39, *SD* = 1.42) and the ACT group (*M* = 1.18, *SD* = 1.57), *t*(10.3) = -1.87, *p* = .090, *d* = -1.05 (see Table 2 for an overview of changes in means of several variables of interest). The ACT group had a higher mean score of changes from pre to post with 1.18 points on the FOSQ indicating an improvement in HRQoL while the MBSR group had a change of -0.39 points. The raise in HRQoL in the ACT group is in line with the reduction in EDS leading to the improvement of HRQoL in the ACT group. Changes in HRQoL from pre to follow-up between the MBSR group (*M* = 0.16, *SD* = 1.20) and the ACT group (*M* = 1.27, *SD* =.89) were also marginally significant, *t*(10.8) = -1.92, *p* = .082, *d* = -1.04. However, neither a follow up dependent paired samples t-test for the MBSR group by itself, *t*(6) = 0.73, *p* = .49 nor for the ACT group reached statistical significance *t*(5) = -1.84, *p* = .13. We found a trend wise significant effect on insomnia symptoms in the MBSR group between pre (*M* = 14.57, *SD* = 4.43) and post intervention (*M* = 12, *SD* = 5.86), *t*(6) = 2.27, *p* = .063, *d* = 0.86 [-0.04, 1.72], and a significant effect in ACT group between pre (*M* = 15.88, *SD* = 3.98) and post (*M* = 15.33, *SD* = 4.46) *t*(5) = 3.50, *p* = .017, *d* = 1.43 [0.23, 2.58]. These findings in both the ACT and MBSR groups are well in line with previous findings showing that they both reduce insomnia symptoms. There was at the same time no significant group difference in the changes of insomnia symptoms (t = -.89, df = 7.65, p = .40), sleep quality (t = -1.64, df = 10.23, p = 0.13) and QoL (t = 1.12, df = 10.06, p = .29). Using the same tests as before for between and within group comparisons, we also found no significant effects on stress, anxious symptoms, and depressive symptoms.

In an additional LMM analysis, concerning the effect of stress, we used daytime sleepiness as the dependent variable and group, stress and their interaction as fixed and intercepts for subjects as random effects. In the model, there was a significant main effect of (ACT) group β = -11.43, *SE* = 3.65, 95% CI [-18.03, -4.72], *t*(17.4) = -3.13, *p* = .0059, a significant main effect of stress β = -0.11, *SE* = 0.04, 95% CI [-.19, -.04], *t*(26.9) = -2.80, *p* = .0093, as well as a significant interaction of stress and group β = 0.16, *SE* = 0.06, 95% CI [.05, .27], *t*(18.8) = 2.73, *p* = .013. In line with this, a likelihood-ratio test for model comparison indicated that the model including the independent variable group provided a better fit for the data than a model without it, χ2(1) = 9.98, *p* = .002. Similarly, a model including the interaction of group and stress provided a better fit than a model without it χ2(1) = 7.90, *p* = .005. However, a likelihood-ratio test comparing a model with stress as an independent variable to one without stress was not significant, χ2(1) = 1.34, *p* = .247.

### Potential other influences

We found neither significant group differences of credibility between the MBSR group (M = -.73, SD = 2.19) and the ACT group (M = .73, SD = 2.98), *t*(11) = 1.05, *p* = .32, nor for expectancy between the MBSR (M = -.67, SD = 2.55) and ACT group (M = .67, SD = 3.12), *t*(11.5) = -.88, *p* = .40. We also found no significant differences in alcohol consumption across all participants between pre (M = 3.21, SD = 4.17) and post intervention (M = 2.77, SD = 4.30), *t*(12) = .61, *p* =.55 and no significant difference across all participants in their BMI between pre (M = 27.58, SD = 4.70) and post intervention (M = 27.76, SD = 4.87), *t*(12) = -.05, *p* = .96. The number of PAP adherent subjects across the time points was rather stable with 69% (eleven participants) before the intervention, 62% (eight participants) at post intervention and 69% (nine participants) at the follow-up time point were PAP-adherent, with dropouts being removed respectively. We found no significant group difference in the number of PAP-adherent subjects between the groups at post-intervention (χ2 = 3.46, p = .06) using ‘N-1’ chi-squared test.

### Subjective impressions

To further develop the programs used in this study based on the experience in this pilot, participants could give anonymous feedback concerning their respective program when filling out the questionnaires. Several participants in both groups responded that they would have preferred in-person sessions. In the MBSR group, many participants remarked that they found the mindful yoga exercises particularly helpful. In the ACT group, some participants wanted exercises that are better tailored to their needs.

The teachers of the respective programs were also asked about their impressions, particularly what they saw as working well or being difficult for participants. In the MBSR group, in line with the participants’ feedback, Yoga exercises seemed to work particularly well. Shorter meditations to recognize one’s own state were also helpful while participants tended to get (more) tired or fall asleep during longer meditations. For the ACT group, the exchange among participants and subsequent discussion, especially regarding subjective experience with the exercises in daily life and individual adaptations of the programs’ content, was important and took up increasingly more time throughout the course.

## 4. Discussion

Despite available medical treatment, there is a high need for studies on behavioral treatments of OSA with rEDS. Our study aimed to start closing this gap by investigating how potential approaches, namely MBSR and ACT, affect symptoms in this patient group and give recommendations for further research. Our results suggest that ACT reduces daytime sleepiness and might improve HRQoL and that both ACT and MBSR reduce insomnia symptoms.

Because we conducted a pilot study with a relatively small sample size, we see the results of our tests as preliminary and in need of further high-quality and well-powered studies.

We found a significant reduction of daytime sleepiness from before to after the intervention in the ACT group. In addition, the change in daytime sleepiness from before to after the intervention differed significantly between the MBSR and ACT group. These are new clinically and statistically significant findings for a behavioral treatment approach of rEDS in OSA patients. They are consistent with one previous finding of a psychotherapeutic approach in a feasibility study (Ong et al., 2020) showing reduced ESS scores in patients with hypersomnia disorders after the intervention. It’s also worth noting the big effect size we found and that the mean reduction of daytime sleepiness in the ACT group was 2.17 points on the ESS, meeting our preregistered criterion for clinical significance of 2 points. The wide 95% CI on the effect size however indicates that further studies are needed to pin down the size of this effect.

It should be mentioned that these effects were not statistically significant anymore at the follow-up time point. The interpretation of this is however difficult. While the mean score of the ACT group is almost unchanged compared to the post time-point (see Table 2) and seems to be relatively stable, the variance increased which might explain the lack of significance. At the same time, however, the mean at the follow-up time point is also influenced by one rather extreme value (see Figure 1). Therefore, it remains unclear whether the EDS reduction of ACT persists over longer periods. Certainly, more research is needed.

One might wonder why, in contrast to our hypothesis and some suggestive previous findings on MBSR, participants in the MBSR group did not show a significant improvement in their EDS. There are several possible explanations. To begin with, the absence of a significant effect in our study cannot be interpreted as showing that mindfulness interventions have no influence on daytime sleepiness. There may be an effect of MBSR on daytime sleepiness in OSA patients with rEDS that we were unable to detect with a rather small sample size. However, with a 95% CI of [-1.78, 3.50] points on the ESS for the mean of change from pre to post intervention, the potential improvement is likely to be smaller than the one we observed in the ACT group. This might be in line with some previous findings where alerting attention, in contrast to orienting and executive attention, was a later development in training MM (Tang et al., 2015). In this case, our results for the MBSR group may indicate that increases in orienting and executive attention alone do not suffice for a clinically significant reduction of EDS. Consistently, it is also plausible that there are either no or rather small effects of MBSR on EDS for OSA patients in the short term, while these potential effects may increase over longer time periods as alerting attention increases. An important qualification of the absence of a significant reduction of EDS in the MBSR-group is that both participants in the MBSR group and the MBSR teacher had the impression throughout the program that mindful yoga was much more helpful than the other components of the program, and thereby potentially effective. Notably, these yoga exercises, which aim to foster an attentive engagement with the present, make up only a relatively small part of the MBSR program. It is possible that participants’ daytime sleepiness may improve in programs using primarily movement-based exercises, i.e., exercises that ensure a basis of alertness in patients with EDS for training mindfulness. If this is the case, it would more naturally cohere with previous research showing improvements of fatigue after training in MM (Xie et al., 2020; Zhang et al., 2019). For further studies, we recommend and encourage the use of such exercises. One further possibility we haven’t investigated here is that either more intensive practice is beneficial or that different kinds of meditation result in bigger changes of EDS.

We also found a marginally significant difference of changes in HRQoL from before to after the intervention between the two groups. The increase in the mean score on the FOSQ in the ACT group was indicative of an improvement of HRQoL. Taken together with the reduction in EDS in the ACT group, it seems plausible that the reduction in EDS led to the improvement of HRQoL in the ACT group. Notably, this difference in changes between the groups remained marginally significant when we compared the pre to follow-up levels of HRQoL between the two groups. At the same time, follow-up tests on HRQoL in the ACT group itself were not significant, which might have been due to the small sample size. It is also possible that ACT itself, independent of EDS led to an increase in HRQoL though it’s noteworthy that the questions in the FOSQ are specific to problems with sleepiness and sleep so that it seems less likely for them to be strongly influenced by general factors of ACT.

We see the marginally significant reduction of insomnia symptoms in the MBSR group as a replication of several earlier findings that found insomnia reducing effects of MM/MBSR (Rash et al., 2019). We also found that ACT significantly reduced insomnia symptoms, and thereby equally replicated newer findings (Paulos-Guarnieri et al., 2022; Salari et al., 2020). It’s worth noting that despite MBSR reducing insomnia symptoms only marginally significantly, the effect size was notably bigger in the MBSR group. Further, the fact that several rather different behavioral therapies, including most prominently CBT-I (not investigated here) are effective in reducing insomnia symptoms, might speak in favor of more general mechanisms for this reduction. Further studies are needed to identify how different behavioral therapies reduce insomnia, thereby potentially uncovering common mechanisms. If there are different mechanisms of these therapies however, they might be helpful for different groups or provide additive effects in severe cases. This possibility would hold additional promise for the treatment of insomnia as well as the common comorbidity of OSA and insomnia.

Our findings regarding stress and sleepiness were mixed. While there was a significant influence of stress in our full LMM, the model comparison using a likelihood-ratio test did not indicate a better fit when including stress as a variable compared to an otherwise identical model. The interaction of stress with group on daytime sleepiness was significant in both cases. However, since the estimated parameter size and its CI are far below one point on the ESS, this is unlikely to be of clinical relevance. Despite this, stress may still be a variable worth including in future studies. This is because it is likely to be interdependent with sleepiness where more stressed individuals tend to be less tired in the short term due to arousing effects, while probably being more exhausted and thereby tired in the long term.

The different reported effects on EDS, HRQoL and insomnia symptoms, are unlikely to be a result of demand or expectancy effects. Because of the employed partial blinding, we expect these factors to be similar in both groups. We did not find significant group differences in either credibility or expectancy as assessed by the CEQ either. Being able to exclude these is an important strength of this study. It’s also unlikely that baseline differences (in PAP use, weight, age, or alcohol consumption) explain later group differences in sleepiness. First, none of them were significantly different between the groups at baseline, presumably due to randomization. Second, we also found no significant changes of these variables throughout the study so that we see it as unlikely that they explain the effects on EDS, HRQoL and insomnia symptoms.

Considering the potential clinical significance and large effect size that we found, it’s important to discuss the potential of behavioral treatments for OSA with rEDS and potentially sleep disorders with EDS more generally. First, for OSA with rEDS, behavioral therapeutic approaches based on ACT seem especially promising given the observed improvement of EDS, HRQoL and insomnia symptoms. ACT might be further adapted to the needs of these patients, especially by addressing common challenges of patients including their symptoms and resulting everyday problems as well as difficulties with the PAP therapy which OSA patients with rEDS are undergoing. Here, therapists might profit especially from knowledge and experience with sleep medicine and a close contact with primary (medical) care providers. Including further established treatment approaches might be another opportunity to reduce patient’s symptoms further. Two potential approaches for this are sleep hygiene and instructions for avoiding supine sleep position since the effectiveness of both is well established. In line with the general approach in ACT, it is at the same time essential to communicate realistic expectations to the patients, especially since the effectiveness of the approaches varies. In total, we see behavioral therapies building on these results as a promising new area of research that deserves more attention, in both OSA patients with rEDS and sleep disorders with EDS more generally.

We further see especially EDS, HRQoL and insomnia symptoms as variables of interest that future large-scale studies should investigate further while ideally incorporating objective measures like the Multiple Sleep Latency Test or apnea-hypopnea index. For future studies, we recommend the use of strong active control conditions as employed here. This is important to single out effects that are specific to the treatment itself while going beyond expectancy and to allow the development of more specific evidence-based therapies in the future. We also encourage the usage of LMMs to deal with missing data more appropriately by reducing sample bias (or estimation bias in the case of imputation of missing values) while making use of all available data points. These models also avoid the assumption that missing data remain stable as the contested approach last-observation-carried-forward (LOCF) does (Blankers et al., 2010) and instead assume that data are missing at random

### Limitations

The lack of objective outcome measures like the latency to fall asleep at daytime from the *Multiple Sleep Latency Test* (MSLT) or the *apnea-hypopnea index* (AHI) from PSG or Home Sleep Apnea Test (HSAT) devices is one limitation of our study. It would be preferable in future studies to include such objective measures which allow going beyond participants’ self-report while enabling a link to subjective measures. Another limitation is the rather small sample size which might have precluded finding some effects while leaving the effect size estimates rather wide. Further, the online setting of the interventions might have curtailed their effectiveness and may have led some people to abstain from participation. The preference for in-person settings might also be connected to the rather high age of our sample and OSA patients generally. Nonetheless, web-based delivery has notable advantages including accessibility and scalability. In our view, it is unclear which approach is more beneficial for OSA patients in total. Consequently, both approaches should be studied further. For further research on behavioral treatment approaches of OSA, it is also desirable to examine different durations beyond the 8-week format investigated here. In our LMMs, we used a limited random effects structure with only subjects as random intercepts since more elaborate random effects structures led to convergence issues, presumably due to the rather small sample size. Finally, we were not able within this study to assess how exactly ACT influences daytime sleepiness, HRQoL and insomnia symptoms nor were we able to assess predictors of effective treatment via ACT. There are many possibilities that should be assessed in future studies.

## 5. Conclusion

Most importantly, we found that ACT reduces EDS for OSA patients with rEDS. This reduction in EDS had a large effect size and is potentially of clinical relevance. We also found that in the ACT-group health-related quality of life improved marginally significantly while insomnia symptoms reduced significantly. Following MBSR, insomnia symptoms also improved marginally significantly, though notably with a bigger effect size. In MBSR, both the MBSR-trainer and participants reported movement-based exercises such as mindful Yoga to be most efficacious. This point may be especially important for studying MM in the context of sleep medicine.

We see our results in total as speaking in favor of ACT as a therapeutic approach for OSA patients with rEDS, thereby being a candidate for an adjunctive therapy. While our study showed the potential of ACT as a therapeutic approach, further high-quality studies are needed to better establish effects and effect sizes, using both subjective and objective measures. Importantly, variations on these ACT-based therapeutic approaches should be investigated. This most importantly includes a more specific adaptation to patients’ needs and might incorporate further components which have already been shown to be efficacious.

## Supporting information

Consort checklist

## Data Availability

All data produced in the present study are available upon reasonable request to the authors

## CRediT author contribution statement

**Max Hellrigel-Holderbaum:** Conceptualization, Data curation, Formal analysis, Methodology, Project administration, Resources, Software, Visualization, Writing – original draft, Writing – review & editing

**Nina Romanczuk-Seiferth:** Investigation, Project administration, Writing – review & editing

**Martin Glos:** Supervision, Writing – review & editing

**Ingo Fietze:** Funding Acquisition, Supervision, Writing – review & editing

## Declaration of competing interest

The authors declare that they have no known competing financial interests or personal relationships that could have appeared to influence the work reported in this paper.

## Acknowledgments

We are grateful to Katrin Lotz-Holz for leading the MBSR group competently and professionally. We also thank Jeehye An, and Benjamín Dupré for discussion and Drs. Meghna Jha, Christian Veauthier, and Maria Kluge for help in participant recruitment.

## Notes

### Competing Interest Statement

The authors have declared no competing interest.

### Clinical Trial

DRKS00026812

### Funding Statement

This study did not receive any funding

### Author Declarations

Ethikkommission of Charite Universitaetsmedizin Berlin, Campus Charite Mitte gave ethical approval for this work

